# Brain volume changes following blast-related mild TBI in service members and veterans: a LIMBIC-CENC study

**DOI:** 10.1101/2024.02.27.24303460

**Authors:** Emily L Dennis, Jared A Rowland, Carrie Esopenko, Nicholas J Tustison, Mary R Newsome, Elizabeth S Hovenden, Brian B Avants, Jessica Gill, Sidney R Hinds, Kimbra Kenney, Hannah M Lindsey, Sarah L Martindale, Mary Jo Pugh, Randall S Scheibel, Pashtun-Poh Shahim, Robert Shih, James R Stone, Maya Troyanskaya, William C Walker, Kent Werner, Gerald E York, David X Cifu, David F Tate, Elisabeth A Wilde

## Abstract

**Importance:** Blast-related mild traumatic brain injuries (bTBI), the “signature injury” of post-9/11 conflicts, are associated with clinically-relevant long-term cognitive, psychological, and behavioral dysfunction and disability; however, the underlying neural mechanisms remain unclear.

**Objective:** To investigate associations between a history of remote bTBI and regional brain volume in a sample of United States (U.S.) Veterans and Active Duty Service Members (VADSM).

**Design:** Prospective case-control study of U.S. VADSM of participants from the Long-term Impact of Military-relevant Brain Injury Consortium - Chronic Effects of Neurotrauma Consortium (LIMBIC-CENC), which enrolled over 1,500 participants at five sites used in this analysis between 2014-2023.

**Setting:** Participants were recruited from Veterans Affairs medical centers across the U.S.

**Participants:** Seven hundred and seventy-four VADSM of the U.S. military met eligibility criteria for this analysis.

**Exposure:** All participants had combat exposure, and 82% had one or more lifetime mild TBIs with variable injury mechanisms.

**Main Outcomes and Measures:** Regional brain volume was calculated using tensor-based morphometry on 3D T1-weighted magnetic resonance imaging scans. TBI history, including history of blast-related injury (bTBI), was assessed by structured clinical interview. Cognitive performance and psychiatric symptoms were assessed with a battery of validated instruments. We hypothesized that regional volume would be smaller in the bTBI group, and that this would be associated with cognitive performance.

**Results:** Individuals with a history of bTBI had smaller brain volumes in several clusters, with the largest centered bilaterally in the superior corona radiata and globus pallidus. Greater volume deficits were associated with a larger number of lifetime bTBIs. Additionally, causal mediation analysis revealed that these volume differences significantly mediated the association between bTBI and performance on measures of working memory and processing speed.

**Conclusions and Relevance:** Our results reveal robust volume differences associated with bTBI. Magnetic resonance elastography atlases reveal that the specific regions affected include the stiffest tissues in the brain, which may underlie their vulnerability to pressure waves from blast exposures. Furthermore, these volume differences significantly mediated the association between bTBI and cognitive function, indicating that this may be a helpful biomarker in tracking outcome after bTBI and suggesting potential treatment targets to prevent or limit chronic dysfunction.

## Introduction

Research on blast-related mild traumatic brain injury (bTBI) has increased exponentially over the last two decades, advancing our understanding of mechanisms and outcomes.^1–5^ Modern warfare, combat training, and advances in weapons technology have exposed service members to blasts at an alarming rate, and the majority of TBIs that occur in war zones include blast as a mechanism of injury.^6,7^ Pre-clinical studies have demonstrated that primary blast forces can directly injure or impair brain structure and function in the absence of other injury mechanisms.^8–10^ As with other TBI mechanisms, clinical outcomes post-blast injury are heterogeneous and only partially explained by injury-related factors. While there are obvious distinctions in mechanism in blast versus blunt/impact TBI (hereon referred to as impact TBI), how distinct the pathophysiological alterations are, both acutely and with long-term cognitive, psychological, and functional outcomes, are less clear. A better understanding of the immediate and long-term effects of bTBI is crucial to improving care and developing targeted interventions for neurobehavioral sequelae.

Advanced neuroimaging has revealed structural alterations that support blast as a unique mechanism of TBI, with white matter (WM) alterations consistently associated with bTBI.^2,11–17^ Although some studies have identified group differences in specific brain regions, many report spatial heterogeneity, indicating that the pattern of disruption varies across individuals.^11,13,16–18^ This inconsistency is likely due to differences in characteristics of each blast exposure including force, direction, additional contemporaneous injury mechanisms (e.g., hit with object, fall from blast energy), and protective factors (e.g., protective gear and physical barriers that partially shield exposure). WM abnormalities have been associated with poor clinical outcomes in several domains including cognition, psychiatric symptoms, and post-concussive symptoms. Additionally, these abnormalities mediate the association between mild TBI (mTBI) and both post-concussive symptoms and cognitive deficits.^16,17^ Veterans with bTBI and memory disruption were found to have metabolite decreases in the hippocampus.^19^ Changes in either brain volume or cortical thickness have also been associated with blast exposure and bTBI. Most studies examining cortical thickness have shown thinning associated with a history of blast exposure and bTBI months and years post-injury,^20–23^ although one study found increased cortical thickness in breachers with repetitive low-intensity blast exposure.^24^ Although fewer studies of brain volume have been published, volumetric changes have been noted in the hippocampus.^3,25^ bTBI has also been shown to alter brain function,^26^ the structure of whole-brain functional connectomes,^27^ functional connectivity throughout the brain,^28^ and functional connectivity within specific circuits.^29,30^ Further, these functional brain imaging changes have also been associated with changes in cognitive performance, specifically working memory.^31^

Together, human and animal studies suggest that bTBI has unique effects on the brain. However, human studies suggest significant individual heterogeneity in blast effects. Novel methods that are robust to this variability are critical for translation to clinical applications. Further, results of structural alterations following bTBI, particularly brain volumes, are sparse and have not been linked to chronic or long-term cognitive outcomes and may suggest that current methods are not sensitive to structural changes following bTBI. Tensor-based morphometry (TBM) involves mapping a participant image to a reference image and using the resulting warp information to calculate voxelwise differences in volume. TBM offers several benefits over other approaches that measure volume, including greater reliability and power in multi-site studies,^32^ and has been applied successfully in mTBI in prior publications, but has not yet been used to study bTBI.^33–35^ FreeSurfer segmentations rely on *a priori* regions of interest (ROIs), whereas the whole brain approach of TBM allows for a data-driven analysis. Voxel-based morphometry (VBM) requires tissue segmentation, which can be problematic in injured brains.

The objective of this study is to evaluate the remote effect of bTBI on brain volume on a voxelwise basis, using TBM in a large cohort of well-characterized veterans and active duty service members (VADSM) with remote bTBI, impact mTBI, and no lifetime TBI, and identify whether alterations are related to cognitive outcomes. We hypothesized that TBM would identify specific areas of the brain most sensitive to the effects of bTBI. Changes in these areas were expected to be related to cognitive outcomes, including performance on tests of attention and memory.

## Materials and Methods

### Participants

This secondary analysis used participant data from the ongoing Long-term Impact of Military-Relevant Brain Injury Consortium - Chronic Effects of Neurotrauma Consortium (LIMBIC-CENC) Prospective Longitudinal Study that is described at length in prior publications.^36–38^ At the time of this dataset extraction, eight sites had participated in data collection. Three sites were excluded from the current analyses due to having too few unexposed control participants for template creation (described below and in further detail in **Supplemental Note 1**). Institutional Review Boards at each site approved this study. All participants provided written informed consent prior to assessment. Inclusion criteria were: 1) prior military combat deployment; 2) combat exposure defined by the Deployment Risk and Resiliency Inventory Section D (DRRI-2-D) score >1 on any item; and 3) at least 18 years of age. Exclusionary criteria were: 1) moderate/severe TBI as defined by standard criteria; or 2) history of major neurologic or psychiatric disorder with significant decrease in functional status and/or loss of ability for independent living (e.g., complete spinal cord injury or schizophrenia).

**Table 1** reports the demographic data of 774 participants that provided usable data (n=670 male, M_age_=40.1, SD=9.8 years).

**Table 1.** Demographic information across sites. For each site included in this analysis, we list the total sample size, the number of males and females, age in years (average and standard deviation), number and percent of participants with a history of any mTBI, deployment-related mTBI, and bTBI, years since last TBI (average and range), years of education (average and standard deviation), active duty status (number and percent), and service branch (percent). Air Force, Army, Marine Corps, and Navy each include their respective Reserves and National Guard divisions.

|  | N (M/F) | Age (avg \| SD) | Any TBI (N \| %) | Deployment-related TBI (N \| %) | Blast-related TBI (N \| %) | Time since last TBI (years, avg \| range) | Education (years, avg \| SD) | Active duty service members | Service branch (%<br>Air Force \| Army \| Marine Corps \| Navy \| No answer) |
| --- | --- | --- | --- | --- | --- | --- | --- | --- | --- |
| Richmond, VA (Site 1) | 252 (207/45) | 44.5 \| 9.6 | 192 \| 76% | 115 \| 46% | 70 \| 31% | 13.4 \| 0.3-51.5 | 14.5 \| 1.6 | 38 \| 15.1% | 8.7 \| 71.8 \| 12.7 \| 6.3 \| 0.4 |
| Houston, TX (Site 2) | 157 (143/14) | 34.0 \| 7.3 | 125 \| 80% | 102 \| 65% | 70 \| 44% | 8.1 \| 0.4-30.4 | 13.4 \| 1.5 | 8 \| 5.1% | 7.6 \| 65.0 \| 18.5 \| 8.3 \| 0.6 |
| Tampa, FL (Site 3) | 205 (177/28) | 38.9 \| 9.2 | 162 \| 79% | 112 \| 55% | 77 \| 38% | 8.7 \| 0.2-50.6 | 14.1 \| 1.6 | 38 \| 18.5% | 12.7 \| 67.3 \| 12.2. \| 7.8 \| 0 |
| Portland, OR (Site 6) | 66 (59/7) | 40 \| 9.5 | 56 \| 85% | 29 \| 44% | 19 \| 29% | 13.1 \| 0.6-38.1 | 14.4 \| 1.6 | 5 \| 7.6% | 16.7 \| 54.5 \| 15.2 \| 13.6 \| 0 |
| Minneapolis, MN (Site 7) | 94 (84/10) | 41.2 \| 10.0 | 63 \| 67% | 33 \| 35% | 16 \| 17% | 16.9 \| 0.5-48.6 | 14.6 \| 1.6 | 15 \| 16% | 9.6 \| 73.4 \| 9.6 \| 7.4 \| 0 |
| Total | 774 (670/104) | 40.1 \| 9.8 | 598 \| 77% | 391 \| 50% | 260 \| 34% | 11.4 \| 0.2-51.5 | 14.2 \| 1.6 | 104 \| 13.4% | 10.3 \| 68.0 \| 13.6 \| 7.9 \| 0.3 |

### Clinical, Neuropsychological, and Emotional Functioning Data Collection

Using a structured interview, lifetime history of all possible concussive events (PCEs) was identified.^39^ PCEs were assessed and classified as mTBI versus not mTBI according to the VA/DoD common definition.^40^. PCE were additionally characterized as occurring during deployment or outside deployment, and as blast-related or non-blast, based on the mechanism of injury. Additional exposures to controlled and uncontrolled blast were also collected.

### Performance and Symptom Validity

The recommended cut-off scores on the Medical Symptom Validity Test and Neurobehavioral Symptom Inventory Validity-10 index were used to detect suboptimal effort on neuropsychological testing and symptom over-reporting, respectively.^41,42^ *Substance Misuse.* Ongoing alcohol consumption within the past three months was evaluated using the Alcohol Use Disorders Identification Test-Concise (AUDIT-C).^43^ *Emotional Functioning*. Assessment of current level of posttraumatic stress disorder (PTSD) symptom severity within the past month was obtained using The PTSD Checklist for DSM-5 (PCL-5) and depressive symptoms within the past two weeks assessed using the Patient Health Questionnaire-9 (PHQ-9).^44,45^ *Cognitive Functioning.* Visual memory and learning were evaluated using the Brief Visuospatial Memory Test-Revised (BVMT-R) Total Recall and Delayed Recall, and verbal memory and learning was assessed using the California Verbal Learning Test-II (CVLT-II) Total, Short, and Long Delay Free Recall scores.^46,47^ Verbal fluency was evaluated using the Delis–Kaplan Executive Function System (D-KEFS) Letter Fluency and Category Switching Total correct variables.^48^ Processing speed was measured using the Wechsler Adult Intelligence Scale 4th Edition (WAIS-IV) Processing Speed Index (PSI), and mental flexibility was assessed using the Trail-Making Test (TMT) -B completion time. A derived score of the completion time of TMT-B-A was used to control for the motor speed element of the test. Working memory was measured using the WAIS-IV Digit Span subtest.^49,50^ Raw scores were used for all cognitive measures because age and gender were included as covariates in analyses. All neuropsychological assessments were completed around the same time as the MRI scan.

### MRI Acquisition

3D T1-W images were collected using a protocol recommended by the Alzheimer’s Disease Neuroimaging Initiative and consistent with other large TBI-based consortia.^51^ All sites implemented monitoring throughout to ensure quality and consistency (e.g., geometric accuracy, signal to noise, adherence to major sequence parameters). See **Supplementary Table 1** for scan parameters.

### Tensor-based Morphometry

A two-step process was used for creating the study-specific template.^52^ Full methods are detailed in **Supplementary Note 1**. Briefly, a site-specific template was created for each of the five sites from 30 unexposed participants without current PTSD, semi-randomly selected to be representative of the overall population. Second, the five site-specific templates were merged into an overall template.

Each participant’s T1-weighted MRI was semi-automatically masked using *antsBrainExtraction* with manual corrections and intensity-normalized with N4.^53^ Resulting files were registered to the site-specific templates using *unbiased_pairwise_registration*.^54,55^ Site-specific templates were registered to the overall template using the same algorithm. The registrations were combined to create a single warp file. The resulting warp files were converted into log Jacobian Determinant files, where positive values indicate larger volumes in the participant image than the template and negative values indicate smaller volumes.

### Statistical Analyses

Voxelwise linear mixed effects models were implemented with *lme* in R3.1.3 with site as a random effect, and age and gender as covariates. Intracranial volume (ICV) was not included as a covariate as the affine and rigid registration steps account for differences in overall brain size. Results were corrected for multiple comparisons correction using searchlight FDR^56^ and reported as Cohen’s *d*.

### Primary Analyses

Our primary group analysis compared individuals with a history of bTBI to those with no history of bTBI (no-bTBI), which included both TBI negative individuals and those with only non-blast mTBI.

### Sensitivity Analyses

We reran the primary comparison with multiple potentially confounding factors included (i.e., PTSD symptoms, depressive symptoms, and/or problematic alcohol use), both as categorical variables using standard cut-off scores and as continuous variables. We additionally re-analyzed covarying for years of education and number of lifetime mTBIs. Within individuals with a history of deployment-related mTBI, we compared the bTBI group to those with impact TBI to separate the blast mechanism from the deployment context. We examined associations between several blast exposure variables and regional volume, including number of bTBIs and number of blast-related PCEs.

### Cognitive Analyses

We examined associations between regional volume and raw scores from the cognitive measures, both across the whole sample and within the bTBI group only. When there was cluster overlap between cognitive performance and results from the primary group comparison, we conducted a modified causal mediation analyses using the R package *mediation*.^57^ For these analyses, the initial predictor was bTBI (yes/no), the mediator was cluster volume from the primary group comparison, and the outcome variable was cognitive performance. In the modified causal approach^58^ the total effect does not need to be significant to proceed with a mediation analysis if theoretical conditions are met, specifically that the effect of the predictor on outcome is not immediate and that mediation analysis offers greater power than separate bivariate tests.

## Results

### Primary Group Analysis

We found multiple areas of smaller volume in the bTBI group compared to no-bTBI, particularly in subcortical GM and central WM (cluster peak Cohen’s *d* range = -0.23 to -0.38; **Figure 1**; **Table 2**). For follow-up analyses, volumes were extracted for these clusters for each participant by taking the average log Jacobian determinant across the cluster mask. The bilateral superior corona radiata (SCR) clusters were merged into a single “SCR” mask, and the bilateral globus pallidus (GP) clusters, which overlapped with nucleus accumbens, substantia nigra, cerebellar peduncles, and internal capsule, were merged into a single “GP” mask.

**Figure 1.**
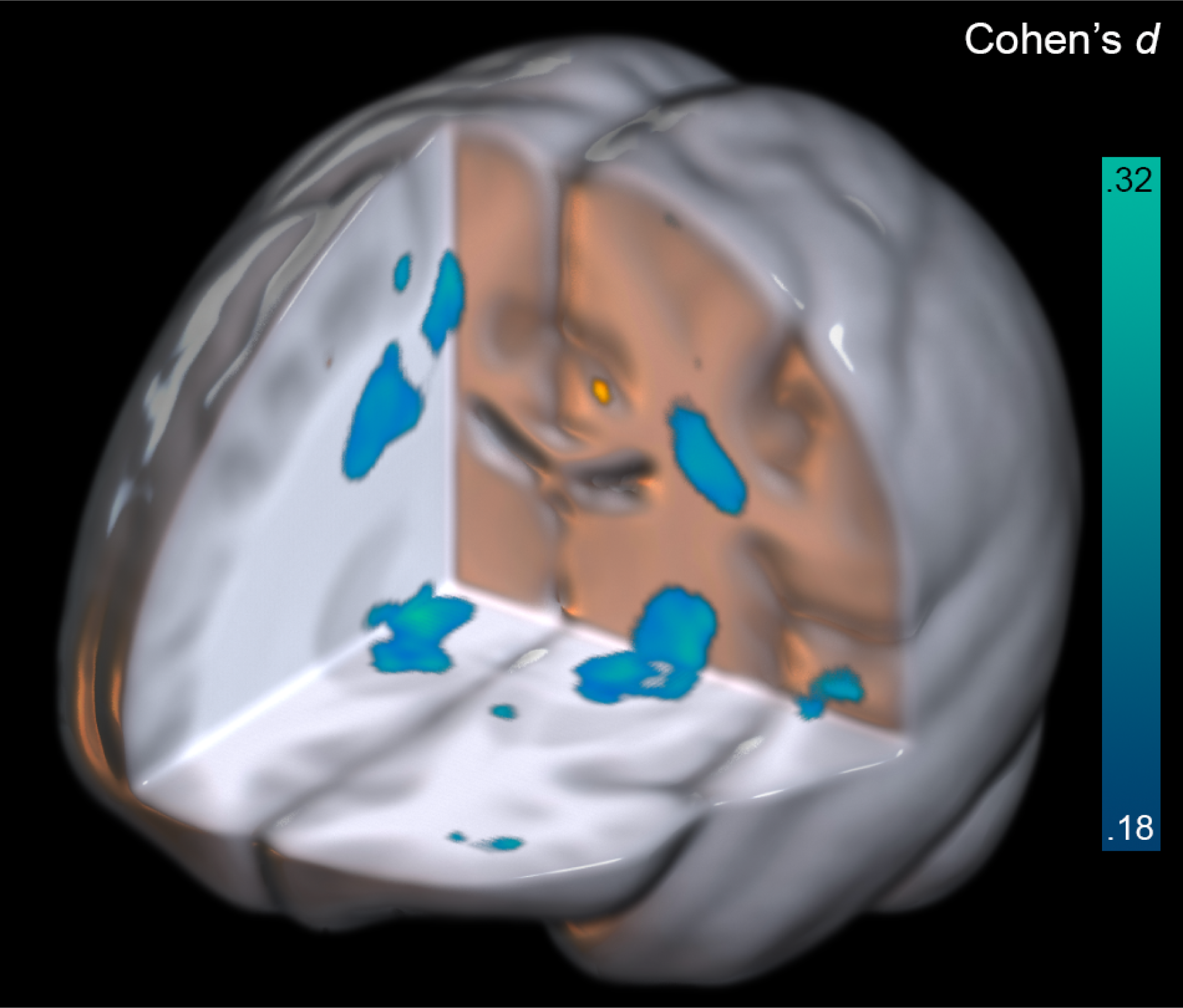
Differences in regional volume between participants with and without a history of bTBI. Clusters showing significantly smaller volumes are shown, with color corresponding to Cohen’s *d*. Left in image is right in brain. Orange clusters indicate larger volumes, most of which are not visible from this view.

**Table 2.** Group differences in volume. Clusters showing significant volumetric differences are shown. For each cluster the cluster size, maximum Cohen’s *d*, MNI coordinates, region, and tissue type are shown. R=right, L=left, WM=white matter, GM = gray matter.

| Positive |  |  |  |  |  |  |  |
| --- | --- | --- | --- | --- | --- | --- | --- |
| Voxels | MAX Cohen's *d* | X | Y | Z | Side | Structure | Tissue |
| 1911 | 0.252 | 18 | -68 | -35 | R | Cerebellum | WM |
| 1425 | 0.315 | 30 | -61 | -2 | R | Lingual gyrus | WM |
| 1371 | 0.38 | 32 | 32 | 6 | R | Inferior frontal gyrus | WM |
| 1305 | 0.278 | -34 | 26 | 1 | L | Insula | GM |
| 934 | 0.27 | -24 | -55 | -3 | L | Lingual gyrus | GM |
| 561 | 0.282 | 56 | -47 | 0 | R | Middle temporal gyrus | WM |
| 528 | 0.263 | -22 | -51 | -48 | L | Cerebellum lobule VIIIB | GM |
| 385 | 0.324 | 29 | 15 | 54 | R | Middle frontal gyrus | GM |
| 336 | 0.274 | 4 | -57 | -35 | R | Cerebellum Vermis IX | GM |
| 269 | 0.303 | 38 | -8 | -20 | R | Middle temporal gyrus | WM |
| 177 | 0.262 | -46 | -63 | 14 | L | Middle temporal gyrus | GM |
| 117 | 0.277 | 29 | 12 | -19 | R | Posterior orbital gyrus | GM |
| 51 | 0.27 | -27 | -63 | 65 | L | Superior parietal lobule | GM |
| 50 | 0.273 | 20 | -89 | 36 | R | Cuneus | GM |
| Negative |  |  |  |  |  |  |  |
| Voxels | MAX Cohen's *d* | X | Y | Z | Side | Structure | Tissue |
| 5300 | 0.311 | 39 | -13 | 67 | R | Superior corona radiata/Precentral gyrus | WM |
| 4397 | 0.294 | -26 | -11 | -3 | L | Globus pallidus/Thalamus/Putamen | GM |
| 3645 | 0.293 | -27 | -10 | 37 | L | Superior corona radiata | WM |
| 2106 | 0.322 | 18 | -3 | -7 | R | Globus pallidus/Thalamus/Putamen | GM |
| 1939 | 0.321 | 36 | 62 | 2 | R | Middle frontal gyrus | GM |
| 1763 | 0.321 | 49 | -70 | -35 | R | Cerebellum Crus I | GM |
| 696 | 0.279 | 2 | -5 | 27 | R | Corpus callosum/cingulum | WM |
| 433 | 0.281 | -29 | -29 | 36 | L | Superior corona radiata | WM |
| 389 | 0.278 | -49 | -16 | -1 | L | Superior temporal gyrus | WM |
| 349 | 0.281 | -1 | -93 | 10 | L | Cuneus | GM |
| 322 | 0.27 | -39 | -80 | -36 | L | Cerebellum Crus I | GM |
| 316 | 0.295 | 42 | 57 | 9 | R | Middle frontal gyrus | GM |
| 303 | 0.262 | 41 | -61 | -52 | R | Cerebellum lobule VII | GM |
| 230 | 0.294 | -7 | 56 | 5 | L | Superior frontal gyrus | GM |
| 211 | 0.25 | -24 | -56 | 32 | L | Superior parietal lobule | WM |
| 203 | 0.256 | -38 | -11 | -43 | L | Inferior temporal gyrus | GM |
| 150 | 0.283 | -37 | 50 | -4 | L | Middle frontal gyrus | WM |
| 124 | 0.263 | -4 | -77 | -23 | L | Cerebellum lobule VII | GM |
| 122 | 0.253 | 62 | 1 | 17 | R | Precentral gyrus | GM |
| 103 | 0.232 | 24 | -23 | 8 | R | Internal capsule | WM |
| 75 | 0.242 | 21 | -1 | 56 | R | Superior frontal gyrus | WM |
| 75 | 0.267 | 37 | -3 | 30 | R | Superior corona radiata | WM |
| 73 | 0.258 | 38 | 49 | 17 | R | Middle frontal gyrus | GM |
| 67 | 0.254 | 52 | 24 | 10 | R | Inferior temporal gyrus | GM |
| 58 | 0.226 | -22 | 49 | 0 | L | Superior frontal gyrus | WM |
| 54 | 0.271 | 28 | -19 | -25 | R | Hippocampal cingulum | WM |

### Sensitivity Analyses

Analyses including additional covariates were largely consistent (cluster peak Cohen’s *d* range = -0.22 to -0.40, **Supplementary** Figure 2). Results remained when restricting analyses to individuals with deployment mTBI, comparing bTBI to impact mTBI, and when covarying for number of lifetime mTBIs (cluster peak Cohen’s *d* range = 0.33 to 0.46).

There was a linear relationship between the number of bTBIs and volume in similar clusters from the primary group analysis (**Supplementary** Figure 3).

### Cognitive Analyses

Across the whole sample, CVLT-II and D-KEFS scores had no significant voxelwise associations so were not considered further. For BVMT-R, both the recall and delayed recall scores were positively associated with regional brain volume (**Supplementary** Figure 4). TMT-B and TMT-B-A completion times were negatively associated with volume (shorter completion time=better performance, associated with larger volume, **Supplementary** Figure 5). WAIS-IV PSI and Digit Span performance were positively associated with volume (**Supplementary** Figure 6). Within the bTBI group, TMT-B, TMT B-A, and Digit Span yielded clusters that were overlapping with the results from the primary group comparison (**Supplementary** Figure 7).

The indirect effects for all six mediation analyses (GP and SCR with TMT-B, TMT B-A, and Digit Span) were significant (*p*s<.05, **Figure 2**).

**Figure 2.**
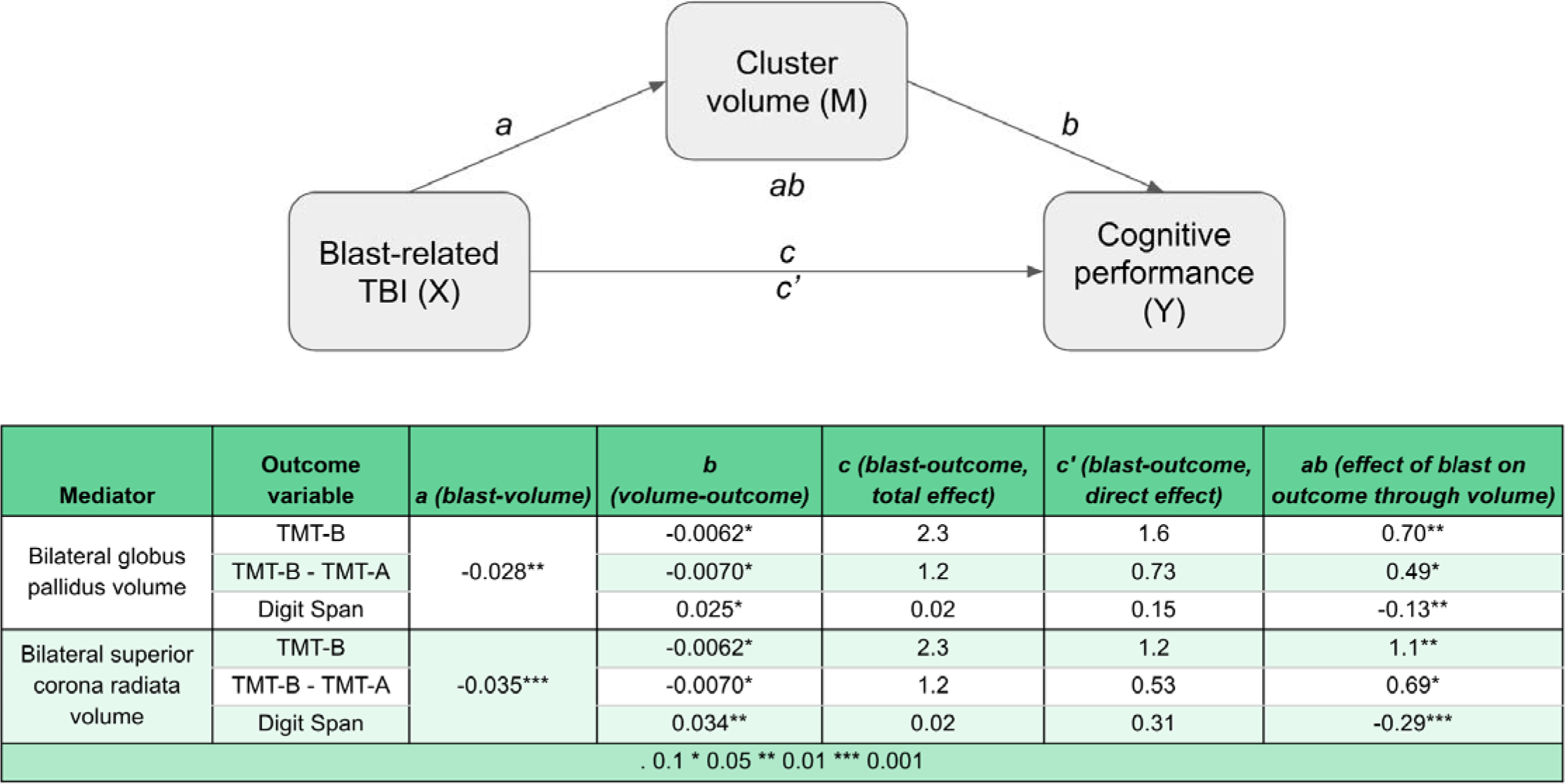
Causal mediation analyses. Results of causal mediation analyses run in R using *mediation* package. For these analyses, the initial predictor was bTBI (yes/no), the mediator was volume of either the superior corona radiata- or globus pallidus-centered clusters (derived as described above in the *Primary Group Analysis* section and depicted in Figure 1), and the outcome variable was TMT-B completion time, TMT-B-A, or Total Digit Span.

## Discussion

We demonstrate that VADSMs with a history of bTBI have reduced volume in WM and subcortical GM regions compared to those without bTBI and that are associated with decreased processing speed and working memory. Reduced volumes remained after adjusting for covariates including PTSD symptoms, depressive symptoms, and substance use. Further, these findings were unique to bTBI, as sensitivity analyses revealed that reduced volumes were not present in VADSMs with a history of deployment-related impact mTBI, and held when controlling for total number of lifetime TBIs. Additionally, a bTBI dose effect was observed, with larger volume alterations among individuals with a higher number of lifetime bTBIs. Overall, these results demonstrate long-term negative outcomes among veterans with a history of remote bTBI (mean=9.8 years, SD=5.0 post-injury). Given the high rate of blast as a mechanism of neurotrauma in modern combat theaters, these results underscore the critical need for continued efforts to mitigate blast-related TBI outcomes even when TBI severity is mild and to identify effective interventions that target bTBI pathology, employ currently available and symptom-based targeted clinical interventions, as well as continued prevention strategies to minimize blast exposure forces in combat and training.

We report smaller volume in the bTBI group in the SCR and in bilateral clusters centered around the GP that included the substantia nigra, nucleus accumbens, internal capsule, and cerebellar peduncle. It is well-established that WM is particularly vulnerable to disruption after TBI.^59^ The shearing forces of TBI stretch and disrupt axons, causing a chemical and physiological imbalance, or axonopathy with greater focal force.^60,61^ Brain tissue stiffness may be another salient outcome of bTBI. Magnetic resonance elastography (MRE) quantifies these characteristics, and studies have shown that the subcortical GM (in particular the pallidum), and WM (in particular projection tracts like the *corona radiata*) have the highest shear stiffness.^62^ Less flexible tissues may be more prone to disruption due to a pressure wave, which may partially underlie the vulnerability of these regions to blast injury.

Several studies have demonstrated cortical thinning in bTBI, primarily in the frontal cortex,^21–23^ but volumetric associations are less common.^3,63^ Our work suggests that there *are* gross alterations in brain volume remotely after bTBI specifically, and contributes to a growing acknowledgement of the unique and adverse long-term effects of blast exposure and bTBI on brain function. Many combat and training missions continue to involve high levels and prolonged exposure to blast waves. Blast exposure is associated not only with a high rate of TBI, but also PTSD, personality changes, cognitive deficits, and suicidal behavior.^64^ Our work adds to these earlier findings by showing measurable changes in brain structure related to differences in cognitive function, unique to bTBI.

MTBI has been linked to changes in cognitive functioning, in particular processing speed and working memory.^65^ However, a neuropathological link between mTBI history and changes in cognitive performance has been less frequently reported. In healthy individuals, the regions implicated in processing speed are lateral frontal and temporal cortices, inferior temporal and parietal cortices, and uncinate fasciculus (connecting frontal and temporal regions).^66^ Working memory performance is similarly supported by prefrontal structures, along with basal ganglia.^67–69^ In mTBI, alterations in WM organization in fronto-thalamic and frontotemporal tracts have been linked to deficits in processing speed and working memory.^70–72^ In bTBI, reduced cortical thickness in frontal regions is associated with poorer performance on an executive function composite score.^22^ We report that changes in volume partially mediated the relationship between bTBI and measures of processing speed and working memory, providing a potential brain-behavior link. Reductions in processing speed performance involving motor output are often associated with dysfunction in the basal ganglia and frontal cortical-subcortical WM tracts. The inherent variability in structural changes at the individual level significantly reduces the likelihood of observing specific functionally localized impairments (e.g., dysphasia, amnesia, lateralized motor deficits). However, the current results cannot rule out that these changes may occur on an individual basis.

Limitations of our study include the retrospective determination of mTBI-related variables which is susceptible to self-report error such as recall bias; however a validated TBI structured interview was utilized to mitigate this as much as possible, provide robust standardization, and avoid clinician-level bias in diagnosis. Second, we did not have sufficient data to parse whether subconcussive blast exposures have an additional effect. The PLS added data collection of the BETS (Blast Exposure Threshold Survey^73^) in 2020 to address this limitation but the current sample size for this analysis was insufficient. Third, the group-based analysis we conducted will only detect abnormalities that are consistent across a group, and thus will miss subject-specific abnormalities.

### Clinical Significance and Future Directions

These results may aid in prognosticating outcomes after injury. By demonstrating the specific cognitive domains that are affected by bTBI, our findings also inform cognitive rehabilitation targets as well as development of proactive interventions to preserve those functions. Further, these results provide evidence regarding the potential underlying causes of the long-term consequences of bTBI and demonstrate that bTBI has unique consequences compared to deployment mTBI generally. Our results indicate that a history of bTBI should be continuously considered in regular medical care and any subsequent neuropsychological evaluations. MRE studies are needed to look specifically at tissue stiffness and determine whether alterations seen in bTBI are indeed related to these tissue properties. Changes in military strategy, protective equipment, and/or artillery design are necessary to limit brain damage in military service members. Clinicians treating VADSMs should not treat a patient’s history of bTBI as a static event but should consider the potential chronic and remote adverse effects on neurobehavioral health over the lifespan.^74^

## Data Availability

Data are available upon reasonable request to the LIMBIC-CENC consortium.

## Acknowledgements

We gratefully acknowledge the Veterans and Active Duty Service Members who participated in this study. This work was supported by the Assistant Secretary of Defense for Health Affairs endorsed by the Department of Defense, through the Psychological Health/Traumatic Brain Injury Research Program LongTerm Impact of Military Relevant Brain Injury Consortium (LIMBIC) Award W81XWH18PH/TBIRPLIMBIC under Awards No. W81XWH1920067 and W81XWH1320095, and by the U.S. Department of Veterans Affairs Awards No. I01 CX002097, I01 CX002096, I01 HX003155, I01 RX003444, I01 RX003443, I01 RX003442, I01 CX001135, I01 CX001246, I01 RX001774, I01 RX 001135, I01 RX 002076, I01 RX 001880, I01 RX 002172, I01 RX 002173, I01 RX 002171, I01 RX 002174, I01 CX001820, and I01 RX 002170. Dr. Pugh was supported by VA Health Services Research and Development Service Research Career Scientist Award, 1 IK6 HX002608; RCS 17-297. The U.S. Army Medical Research Acquisition Activity, 839 Chandler Street, Fort Detrick MD 217025014 is the awarding and administering acquisition office. Opinions, interpretations, conclusions and recommendations are those of the author and are not necessarily endorsed by the Department of Defense, Department of Veterans Affairs, Defense Health Agency, Uniformed Services University, or Federal Government.

