## Supplementary Materials for "Brain volume changes following blast-related mild TBI in service members and veterans: a LIMBIC-CENC study"

**Supplemental Note 1. Tensor-based morphometry methods.**

**Supplemental Figure 1. Workflow for multi-site template creation (A) and multi-stage registration (B)**. In panel A, the workflow is shown for creating site-specific templates first (along with information about participant selection for templates) and then an overall template using *antsMultivariateTemplateConstruction2*. A total of 150 unexposed (no TBI) participants from 5 sites were used for the overall template. In panel B, the TBM workflow is shown, including registration between the participant images and site templates, and between the site templates and overall template using *unbiased_pairwise_registration*.

**Supplemental Table 1. Scan parameters for each site.**

**Supplemental Figure 2. Overlapping clusters for sensitivity analyses.** Shown are the overlapping clusters from 6 sensitivity analyses: 1) covarying for PTSD (binary), 2) covarying for depression (binary), 3) covarying for problematic alcohol use, 4) covarying for education, 5) blast-related vs. blunt deployment mTBI, and 6) covarying for the lifetime number of mTBIs (of any mechanism or context). Color corresponds to the number of analyses that had significant results (after voxelwise corrections for multiple comparisons) for the given cluster. Axial slices are noted in MNI coordinates. Left in image is right in brain.

**Supplemental Figure 3. Voxelwise associations with number of blast-related TBIs, within the blast TBI group.** Regions whose volume was significantly associated with the number of blast-related mTBIs individuals sustained over their lifetimes are shown. Color corresponds to the *T*-statistic of significant voxels, as indicated in the color bar. Axial slices are noted in MNI coordinates. Left in image is right in brain.

**Supplemental Figure 4. Voxelwise associations with BVMT-R performance across the whole sample.** Regions where volume was associated with BVMT-R Recall and BVMT-R Delayed Recall performance across the whole sample. Color corresponds to the *T*-statistic of significant voxels, as indicated in the color bar. Axial slices are noted in MNI coordinates. Left in image is right in brain.

**Supplemental Figure 5. Voxelwise associations with Trails performance across the whole sample.** Regions where volume was associated with Trails B (with and without controlling for Trails A) completion time across the whole sample. Color corresponds to the *T*-statistic of significant voxels, as indicated in the color bar. Axial slices are noted in MNI coordinates. Left in image is right in brain.

**Supplemental Figure 6. Voxelwise associations with WAIS-IV performance across the whole sample.** Regions where volume was associated with WAIS-IV Total Digit Span and Processing Speed Index performance across the whole sample. Color corresponds to the *T*-statistic of significant voxels, as indicated in the color bar. Axial slices are noted in MNI coordinates. Left in image is right in brain.

**Supplemental Figure 7. Voxelwise associations with cognitive performance in the blast mTBI group.** Regions where volume was associated with WAIS-IV Total Digit Span performance and Trails B completion time (with and without corrections for Trails A) across the blast mTBI group only. Color corresponds to the *T*-statistic of significant voxels, as indicated in the color bar. Axial slices are noted in MNI coordinates. Left in image is right in brain.

**Supplemental Note 1. Tensor-based morphometry methods.**

Studies have shown that a study specific template can improve registration quality.^1^ To do this for our multi-site study, we created the template in two stages, first with site-specific templates and then with an overall template created from the site templates. To create the site templates, we selected 30 unexposed individuals from each site. These were semi-randomly selected to be representative of the age distribution and gender distribution of the overall sample. Three of the initial eight sites did not have enough unexposed participants to create a site template so they were not included in the analysis. For four of the five remaining sites, 28 males and 2 females were selected; one site only had 27 unexposed males, so 3 unexposed females were selected for that site. The site-wise details, along with a schematic for the template procedure, may be found in **Supplemental Figure 1.** All processing was done using ANTs functions (Advanced Normalization Tools):

- *antsMultivariateTemplateConstruction2* for both the site-specific and overall templates
- *unbiased_pairwise_registration* for registering participant images to the site templates and for registering site templates to the overall template
- *antsApplyTransforms* to combine the two registration steps (participant to site template + site template to overall template) to generate a participant to overall template warp file
- *CreateJacobianDeterminantImage* to calculate the log Jacobian Determinant files (logJac) from the warp file

In the resulting files, a logJac value greater than 0 appears white and indicates an area with larger volume than the template image, while a logJac value less than 0 appears dark and indicates an area with smaller volume than the template image.

**Supplemental Figure 1. Workflow for multi-site template creation (A) and multi-stage registration (B)**. In panel A, the workflow is shown for creating site-specific templates first (along with information about participant selection for templates) and then an overall template using *antsMultivariateTemplateConstruction2*. A total of 150 unexposed (no TBI) participants from 5 sites were used for the overall template. In panel B, the TBM workflow is shown, including registration between the participant images and site templates, and between the site templates and overall template using *unbiased_pairwise_registration*.

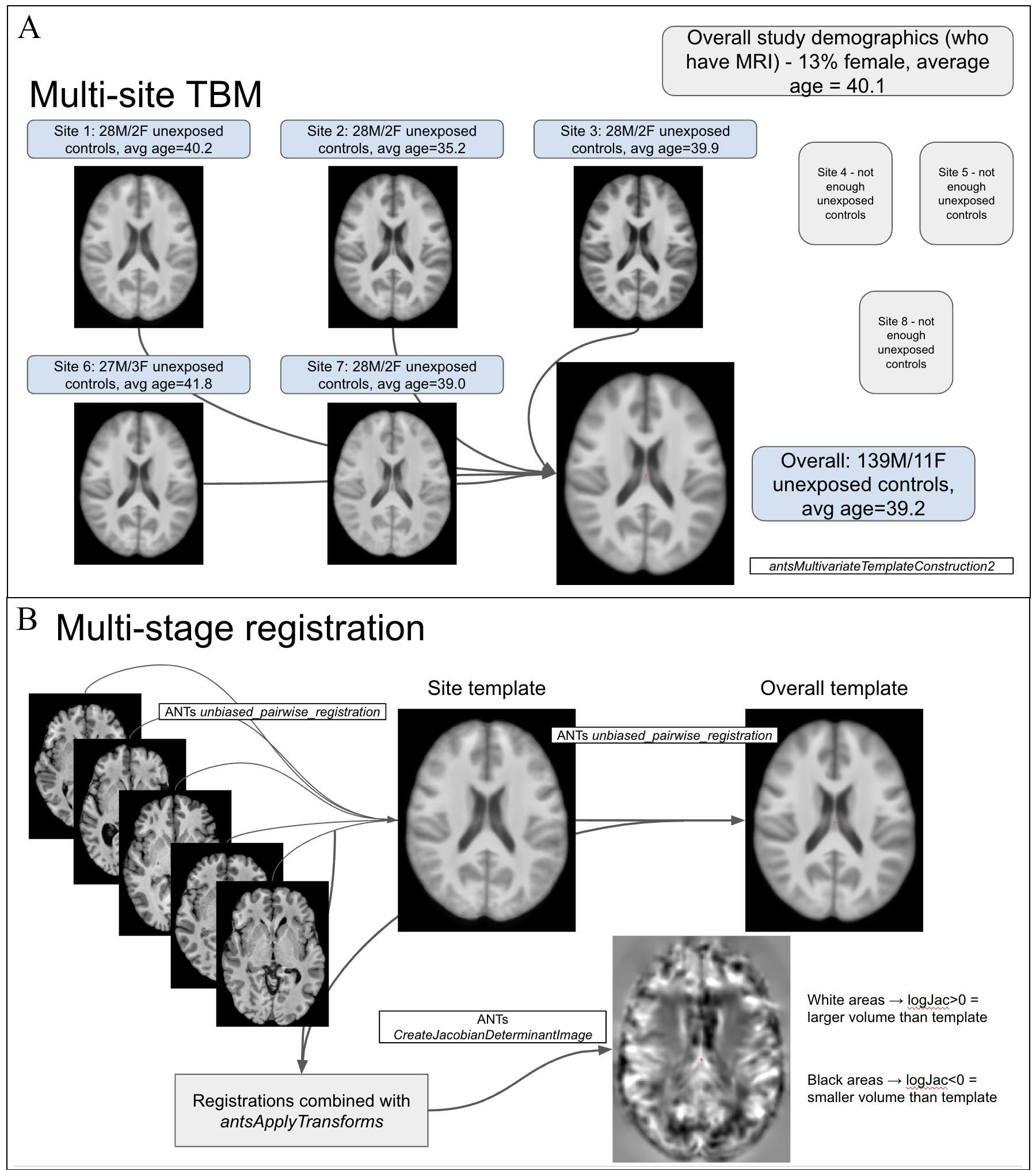

**Supplemental Table 1. Scan parameters for each site.** T1-weighted scan protocols across the six data collection sites included in this analysis. Scanner, TR, TE, voxel size, matrix, and slices are listed for each site.

| **Site** | **Scanner** | **TR (ms)** | **TE (ms)** | **Voxel size (mm)** | **Matrix** | **Slices** |
| --- | --- | --- | --- | --- | --- | --- |
| Richmond, VA (Site 1) | Philips Ingenia | 6.78 | 3.157 | 1x1x1.2 | 256x256 | 176 |
| Houston, TX (Site 2) | Siemens TrioTim | 2300 | 2.96 | 1x1x1.2 | 240x256 | 176 |
| Tampa, FL (Site 3) | GE Signa HDxt | 6.28 | 2.78 | 1x1x1.2 | 256x256 | 196 |
| Portland, OR (Site 6) | Philips Achieva | 6.76 | 3.145 | 1x1x1.2 | 256x256 | 170 |
| Minneapolis, MN (Site 7) | Siemens Prisma | 2400 | 2.24 | 0.8x0.8x0.8 | 300x320 | 208 |

**Supplemental Figure 2. Overlapping clusters for sensitivity analyses.** Shown are the overlapping clusters from 6 sensitivity analyses: 1) covarying for PTSD (binary), 2) covarying for depression (binary), 3) covarying for problematic alcohol use, 4) covarying for education, 5) blast-related vs. blunt deployment mTBI, and 6) covarying for the lifetime number of mTBIs (of any mechanism or context). Color corresponds to the number of analyses that had significant results (after voxelwise corrections for multiple comparisons) for the given cluster. Axial slices are noted in MNI coordinates. Left in image is right in brain.

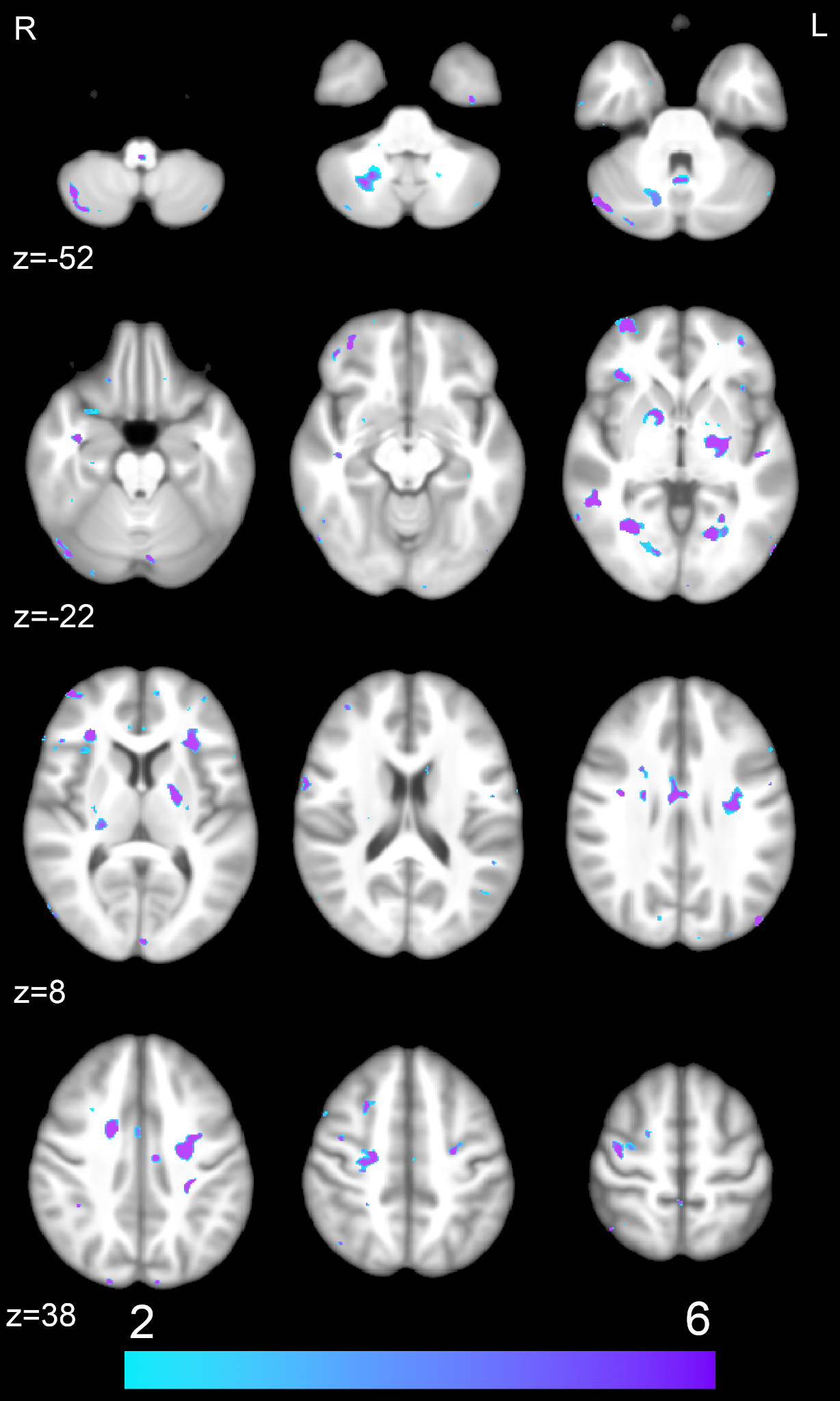

**Supplemental Figure 3. Voxelwise associations with number of blast-related TBIs, within the blast TBI group.** Regions whose volume was significantly associated with the number of blast-related mTBIs individuals sustained over their lifetimes are shown. Color corresponds to the *T*-statistic of significant voxels, as indicated in the color bar. Axial slices are noted in MNI coordinates. Left in image is right in brain.

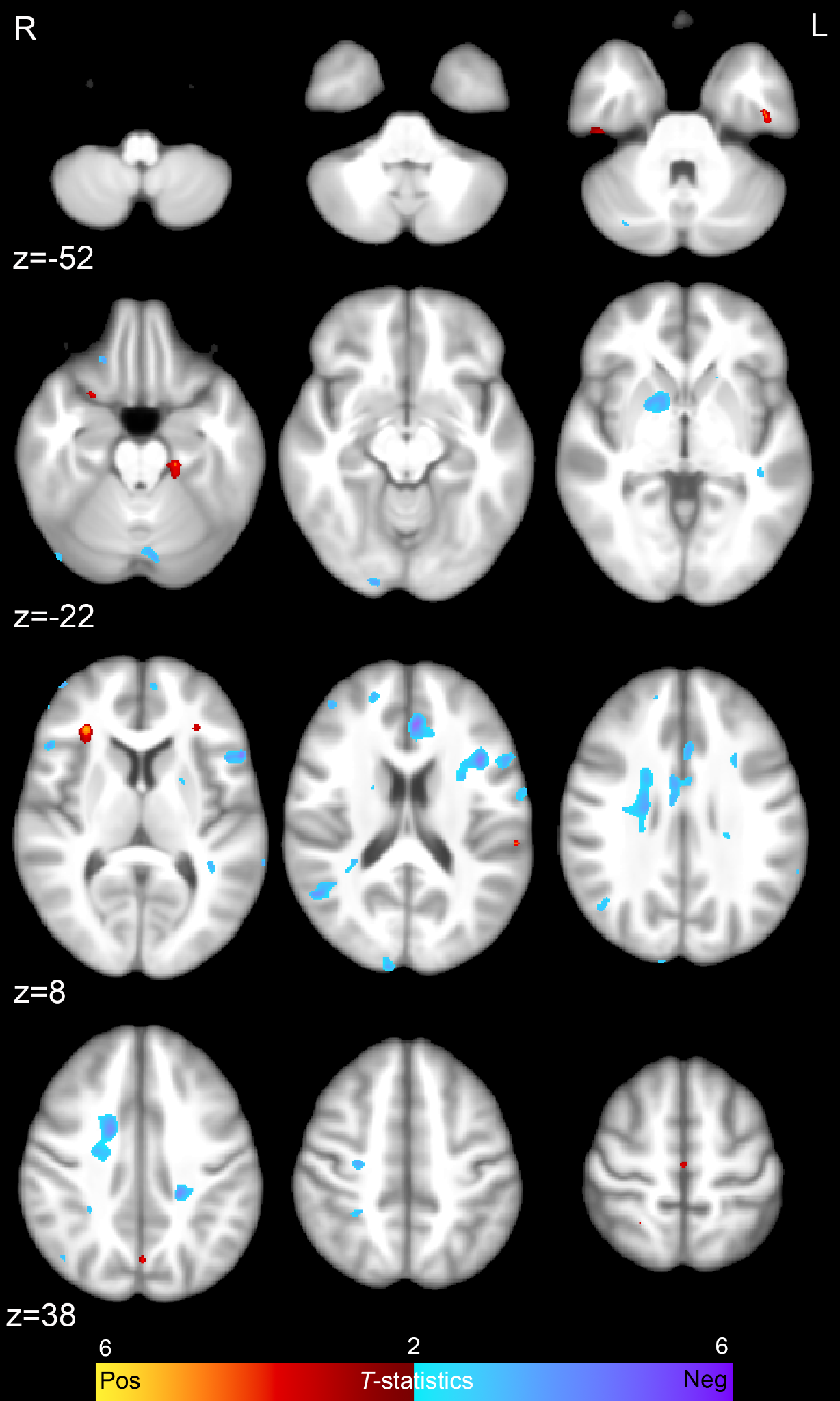

**Supplemental Figure 4. Voxelwise associations with BVMT-R performance across the whole sample.** Regions where volume was associated with BVMT-R Recall and BVMT-R Delayed Recall performance across the whole sample. Color corresponds to the *T*-statistic of significant voxels, as indicated in the color bar. Axial slices are noted in MNI coordinates. Left in image is right in brain.

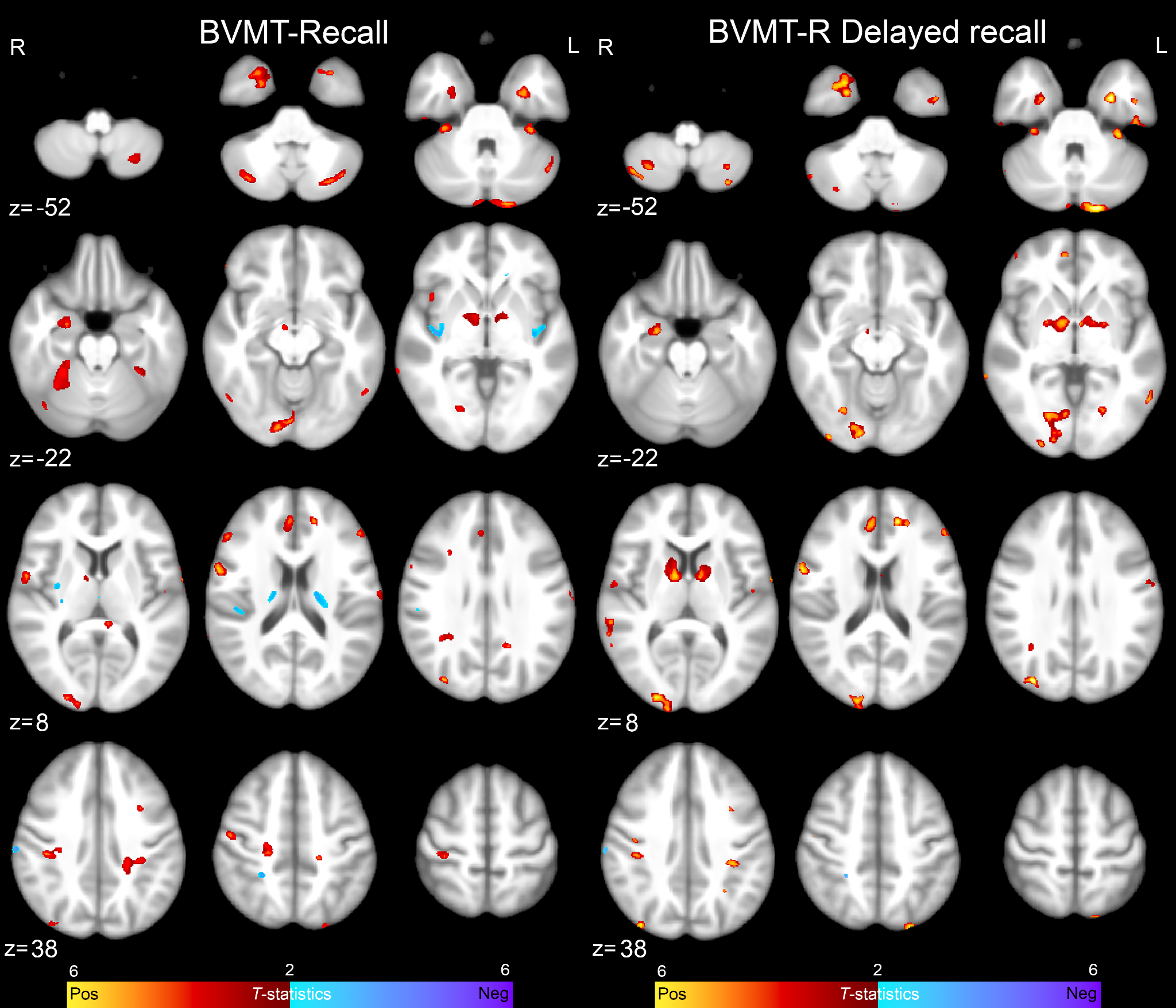

**Supplemental Figure 5. Voxelwise associations with Trails performance across the whole sample.** Regions where volume was associated with Trails B (with and without controlling for Trails A) completion time across the whole sample. Color corresponds to the *T*-statistic of significant voxels, as indicated in the color bar. Axial slices are noted in MNI coordinates. Left in image is right in brain.

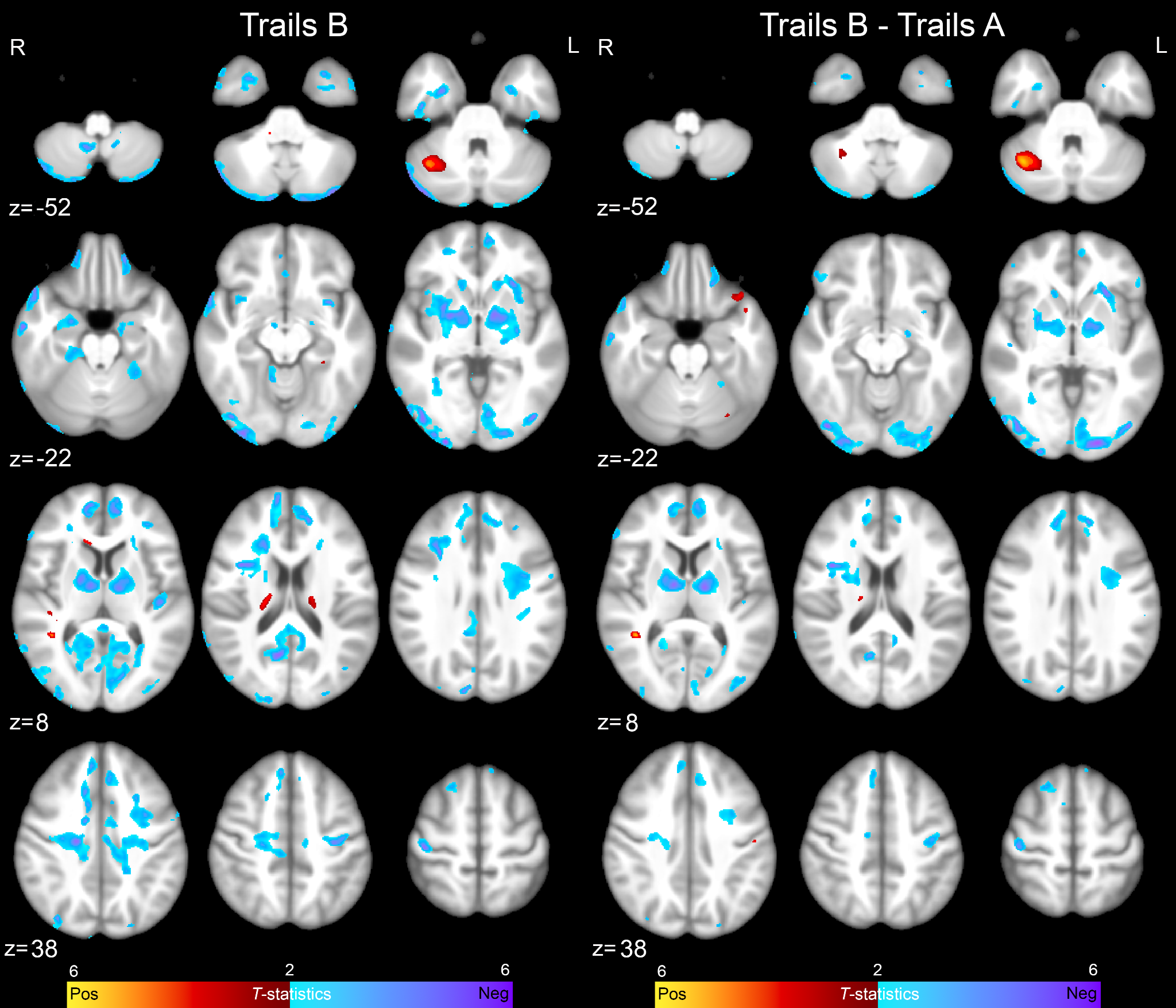

**Supplemental Figure 6. Voxelwise associations with WAIS-IV performance across the whole sample.** Regions where volume was associated with WAIS-IV Total Digit Span and Processing Speed Index performance across the whole sample. Color corresponds to the *T*-statistic of significant voxels, as indicated in the color bar. Axial slices are noted in MNI coordinates. Left in image is right in brain.

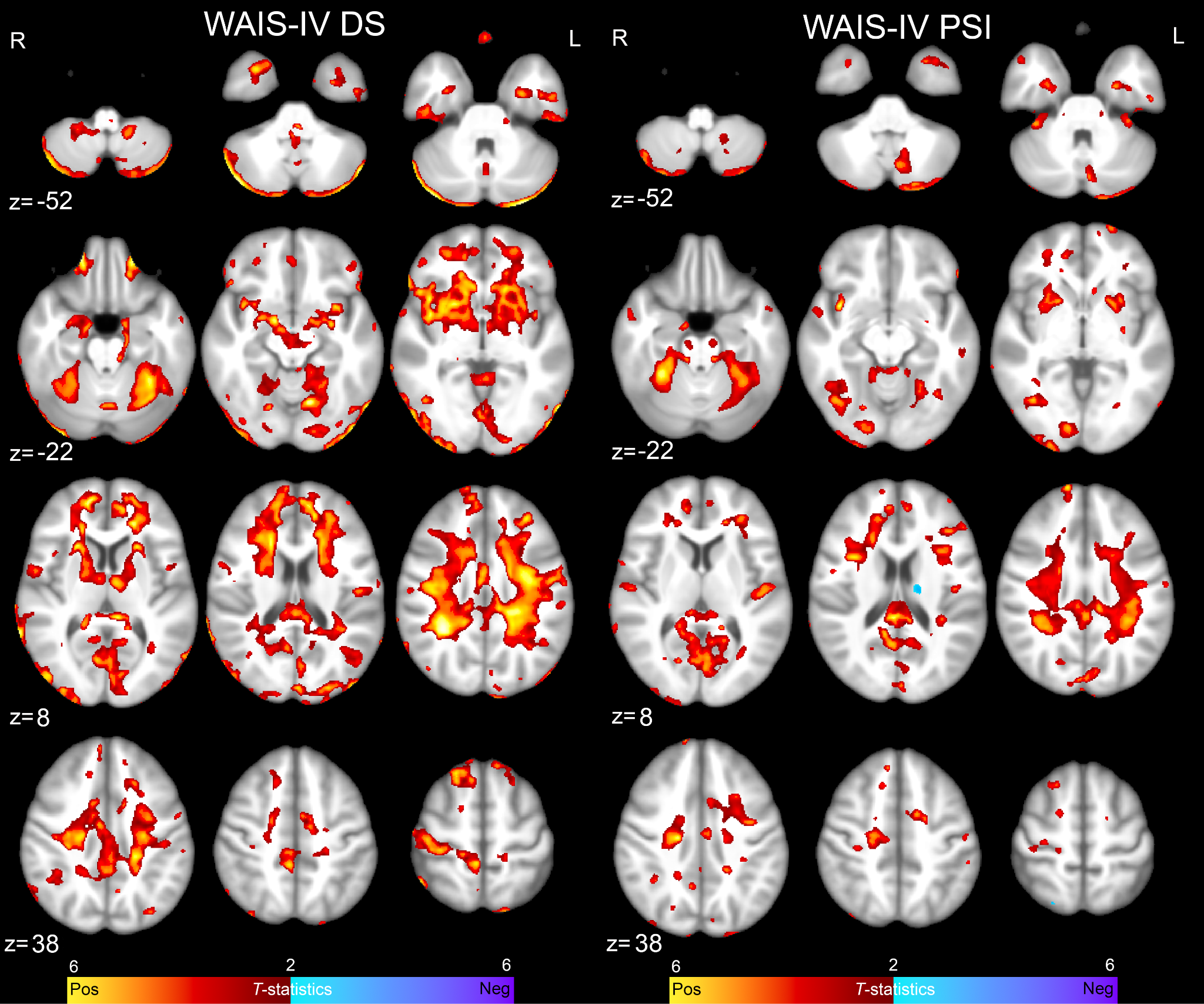

**Supplemental Figure 7. Voxelwise associations with cognitive performance in the blast mTBI group.** Regions where volume was associated with WAIS-IV Total Digit Span performance and Trails B completion time (with and without corrections for Trails A) across the blast mTBI group only. Color corresponds to the *T*-statistic of significant voxels, as indicated in the color bar. Axial slices are noted in MNI coordinates. Left in image is right in brain.

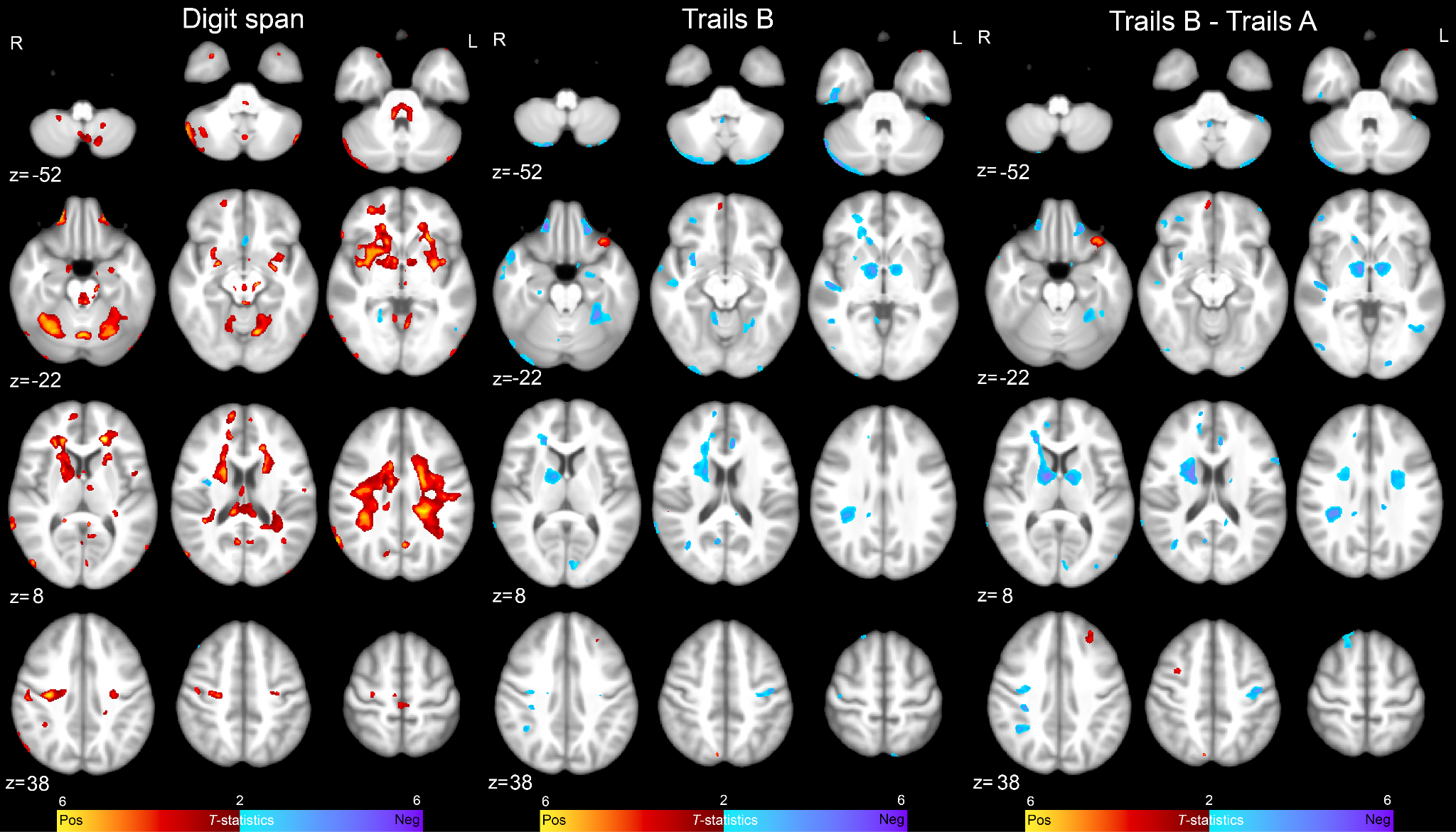
